# TENS for Movement-Evoked Pain in Fibromyalgia: A Randomised Double-Blind Controlled Trial

**DOI:** 10.64898/2026.09.15.26363019

**Authors:** Mathil Ruel, Shayana Lussier Crevier, Nicolas Dehors, Serge Marchand, Isabelle J. Dionne, Guillaume Léonard

**Author notes:** Corresponding author: Guillaume Léonard, P.T., Ph.D., Centre de recherche sur le vieillissement CSSS-IUGS, 1036, rue Belvédère Sud, Sherbrooke (Québec) J1H 4C4.

## Abstract

**Purpose:** Fibromyalgia involves chronic widespread pain and heightened movement-evoked pain (MEP), which can limit access to resistance training’s functional and psychological benefits by hindering exercise participation and adherence. This study aimed to determine whether transcutaneous electrical nerve stimulation (TENS) applied during resistance exercise reduces MEP in women with fibromyalgia.

**Materials and methods:** 21 women with fibromyalgia performed 2 sets of 10 leg press repetitions at 60% predicted 1-RM while receiving randomized active TENS (100 Hz) or sham TENS. Pain intensity and unpleasantness (0–10 numerical rating scales) were assessed at baseline, immediately post-exercise, 15min post-exercise, end-of-day, and 24-h later.

**Results:** Active TENS led to a greater reduction in post-exercise pain than sham, with 90% of participants experiencing at least a 15% decrease in whole-body pain 15 minutes post-exercise, compared to none in the sham group. No group differences were observed at the end of the day or 24h after the exercise session.

**Conclusion:** These results suggest that a single session of high-frequency TENS during resistance exercise can produce short-term reductions in MEP in women with fibromyalgia. While effects did not persist at 24h, these short-term effects may help mitigate acute exercise-related pain and support early training adherence.

## Introduction

Fibromyalgia is a debilitating condition with a complex and multifactorial nature (1). Characterized by persistent widespread pain and a heightened pain sensitivity (2), fibromyalgia disproportionately affects women with a rise in it’s prevalence after the age of 40 (3). Despite advances in pharmacologic treatments, managing fibromyalgia remains challenging, as medications often provide limited symptom relief and carries risk of side effects (4,5).

Beyond pharmacological approaches, physical exercise has emerged as a central pillar in the management of fibromyalgia, supported by decades of research and endorsed as a first-line treatment by the European League Against Rheumatism (4). To this day aerobic exercise is the most studied modality (6). Emerging evidence supports the efficacy of resistance training in reducing fibromyalgia-related symptoms, and improving functional capacity (7). However, these benefits typically emerge gradually and may take up to 13 weeks to be observed (8). In the meantime, the persistence of pain – which is often temporarily exacerbated during and after exercises – remains a significant barrier to exercise (9). A significant number of patients with fibromyalgia report increased pain sensitivity during or after exercise, a phenomenon called exercise-induced hyperalgesia or movement-evoked pain (MEP) (10). Although, MEP shares some similarities with delayed-onset muscle soreness, their distinctive mechanisms, timing, and clinical presentation have been outlined by Butera et al. and other studies (10,11).

Transcutaneous electrical nerve stimulation (TENS) could offer a promising, non-invasive and safe (12) solution to MEP and facilitate adherence to exercise in people with fibromyalgia. By delivering high-frequency (∼100 Hz), low-intensity currents through skin electrodes, conventional TENS modulates pain through both spinal and supraspinal mechanisms (13,14). To our knowledge only two studies have examined the effect of TENS on MEP (15,16), both reporting decreased pain intensity following functional assessments such as the 6-minutes walk test (15,16) and the 5-sit-to-stand test (16). While these functional tasks provide valuable insights, they mainly capture global whole-body performance and do not adequately reflect the specific context and demands of therapeutic exercise. Accordingly, it remains unclear whether TENS exerts similar effects on MEP during targeted, localized mechanical loading such as resistance exercise.

The present study addresses this knowledge gap by investigating whether a single session of conventional TENS, applied before and during resistance training, can reduce short term (<24 h) MEP pain intensity and unpleasantness in women over 40 living with fibromyalgia compared to sham TENS.

## Materials and Methods

### 1.1. Trial Design

This was a double-blind, randomized controlled, inter-subject (independent group) trial with 2 parallel arms (active TENS vs. sham TENS) and minimisation allocation. The study was conducted at the Research Center on Aging (Sherbrooke, Québec, Canada). The trial was registered at ClinicalTrials.gov (<u>NCT06834308</u>, 02/12/2025). Minor protocol modifications, including adjustments to the eligibility criteria, were documented on ClinicalTrials.gov and approved by the local ethics committee, and are detailed in the sections below.

### 1.2. Participants

Twenty-one women living with fibromyalgia aged between 46 and 74 years (mean age: 60±8.5 years) participated in this study. Participants were all French-speaking, community-dwelling individuals.

#### 1.2.1. Eligibility Criteria

Inclusion criteria were: age 40 years or older; diagnosis of fibromyalgia established by a licensed physician; persistent pain in the lumbo-pelvic or lower limb region with an intensity ≥3/10 (0=no pain, 10=worst pain imaginable); refraining from caffeine and analgesics 6 hours before the experiment; and refraining from smoking 2 hours before the experiment. Exclusion criteria included poorly controlled cardiovascular diseases; contraindications to physical activity or to TENS (17,18); prior participation in TENS research or use of a TENS device in the past 10 years; and being physically active prior to the study (i.e. meeting recommendations of 150 minutes per week of moderate to vigorous activity or two sessions of 30 minutes of muscle-strengthening exercises per week for at least 2 months). This last criterion was intended to ensure a relatively inactive participant sample, thereby limiting prior exercise adaptation from confounding the assessment of MEP.

#### 1.2.2. Recruitment and Pre-Visit Assessment

Participants were recruited through local advertisements (e.g., pharmacy, hospital, physical therapy clinics, university campus, fibromyalgia association) or via the Research Center on Aging recruitment bank, with the intention of broadening outreach and enhancing diversity in the study sample. Eligibility was first assessed during a telephone screening interview followed by completion of online pre-visit questionnaires. A sociodemographic questionnaire, including age, gender, income, work status and medication use, was used to describe the study population. Fibromyalgia severity was measured with the Fibromyalgia Impact Questionnaire (FIQ), and kinesiophobia with the Tampa Scale of Kinesiophobia (TSK-17). Physical activity domains (transportation, recreation, and occupational or household) were assessed with the French version of the Physical Activity Adult Questionnaire (PAAQ). Because the PAAQ classifies any ≥10 minute activity accompanied by even mild shortness of breath as moderate intensity, it may lead to an overestimation of true fitness levels, particularly among individuals with asthma (19). Hence, in a few cases where PAAQ scores exceeded the exclusion threshold despite lower reported activity, participants were retained.

During recruitment, eligibility criteria were slightly modified to improve enrollment, including lowering the minimum age requirement (from 50 to 40 years) and removing the menopausal status condition.

### 1.3. Intervention and Comparator

Eligibility criteria regarding contraindication to TENS and exercise were reconfirmed at the beginning of the experimental visit. Both TENS conditions incorporated identical exercise and device usage protocols (see below), differing only in whether active or sham electrical stimulation was delivered. All experimental visits were conducted between 10-am and 3-pm, a time frame generally associated with lower pain symptoms in fibromyalgia patients (20).

#### 1.3.1. Exercise Testing and Protocol

The exercise intervention is reported in accordance with the Consensus on Exercise Reporting Template guidelines (21) to ensure comprehensive and reproducibility. The resistance training protocol consisted of a seated leg press exercise (see figure S1). All participants completed a familiarization period and a warm-up, followed by a repetitions-to-failure (RTF) test to determine their 5-to-10-repetition maximum (RM). The RTF test was conducted according to the American College of Sports Medicine recommendations, allowing up to 3 attempts (22). The knee angle on the leg press was standardized to 90-degrees; due to anatomical considerations such as abdominal pressure or inability to reach 90-degrees, 6 participants performed the exercise at a 120-degree knee angle. The third attempt was considered the final test, regardless of whether the participants exceeded 10 repetitions before reaching muscle failure.

The Epley (1985) equation was used to estimate the one-repetition maximum (1-RM) (23,24):

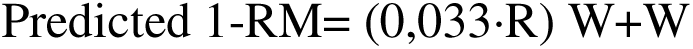

Where R is the number of repetitions and W is the maximal weight lifted. This equation demonstrates excellent validity for inactive older women (24).

In the same visit, participants then performed 2 sets of 10 repetitions at 60% of their estimated 1RM, with a 1–2-minute rest between sets (22). The testing and training session were performed individually and supervised by a kinesiologist.

#### 1.3.2. Active TENS Condition

Conventional high-frequency TENS was administered using a 2-channel TENS unit (Soul Electromedic™) and carbon electrodes. Electrodes were placed on the area identified by the participant as most painful (loco dolenti) (25) in the lumbo-pelvic and/or lower limb region. TENS was applied at 100 Hz and 60 µs, with intensity adjusted to produce strong but comfortable (below pain threshold) paresthesia sensations (26). TENS was applied for 10 minutes before the first set of leg press and then maintained for another 20 minutes during the 2 leg press sets, for a total duration of 30 minutes.

After the experimental visit, participants took the TENS device home for 24-hours. They were instructed to use it as much as they wanted, depending on their pain symptoms. Participants were provided with 2 separate digital logbooks: the first to be completed at the end of the day after the experimental visit, and the second to be completed 24 hours post-visit. Both logbooks were used to record device usage, pain outcomes (see section 1.4) as well as caffeine consumption during their respective periods.

#### 1.3.3. Sham TENS Condition

An identical TENS unit was used in the sham condition; however, stimulation was disabled through the use of modified, non-conductive cables, ensuring no electrical output was delivered. Sham TENS intensity was set to 4 mA to display consistent output on the device screen, despite stimulation being blocked by the modified non-conductive cables. As for the active TENS condition, sham TENS electrodes were positioned over the participant-identified area of greatest pain, and sham stimulation was applied for 10 minutes prior to and during resistance exercise (2-sets of 10-repetitions at 60% 1-RM leg press) for a total duration of 30 minutes. Participants also brought the sham TENS apparatus home, with the same instructions, and were required to record its use, as well as caffeine consumption and pain outcomes, in the 2 digital logbooks.

The TENS interventions were delivered by 2 trained operators (one female [n=11], one male [n=10]). Both followed the same standardized procedures to ensure consistency in intervention delivery and participant interaction. The outcome assessor (MR) remained the same throughout the study.

### 1.4. Outcomes

Whole body (WB) pain intensity (primary outcome) and pain unpleasantness (secondary outcome) were assessed using two separate 0-10 numerical rating scale (NRS, pain intensity: 0=no pain, 10=unbearable pain; pain unpleasantness: 0=not unpleasant, 10=extremely unpleasant). Additional pain intensity and unpleasantness scales were introduced after the first 5 participants to specifically assess the effect of TENS on the lower body (LB), allowing for more precise pain localization in the remaining 16 participants.

All pain outcomes (WB and LB; pain intensity and unpleasantness) were assessed 5 times: at baseline (T1), immediately post-exercise (T2), 15 minutes post-exercise (T3), at the end of the day of the visit (T4), and 24 hours post-exercise (T5). Pain ratings at T1, T2, and T3 were collected in person at the laboratory, while pain assessments for T4 and T5 were completed via participants’ digital logbooks (see overview in figure 1).

**Figure 1.**
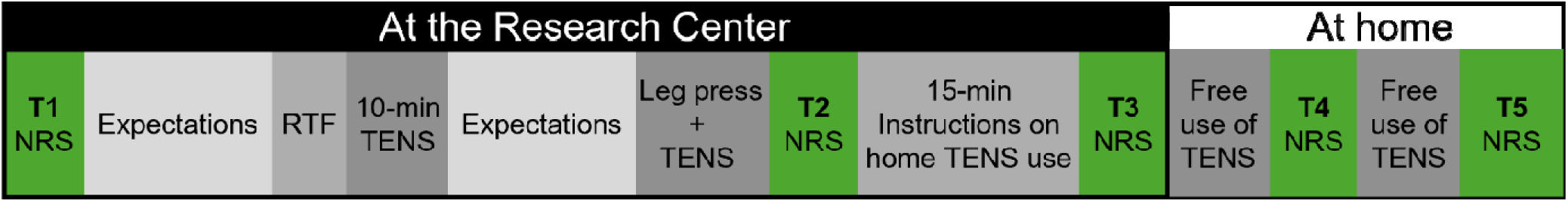
Overview of study procedures. T1 baseline; T2 immediately post-exercise; T3 15 minutes post-exercise; T4 end of the day following visit; T5 24 hours post-exercise; NRS 0-10 numerical rating scale

Participants were monitored for any potential adverse events during exercise and TENS application through spontaneous reporting by participants and study staff.

### 1.5. Sample Size

Sample size was calculated a priori with G*Power (version 3.1.9.7), with a standardised power (0.80) and alpha error (0.05) based on the primary pain outcomes (change scores of WB pain intensity). In the absence of closely related prior studies, a medium effect size (f=0.25) was selected according to Cohen’s widely used conventions (27), representing a difference likely to be clinically relevant (28). As such, 22 participants were estimated to be required, and 24 were recruited to account for possible dropouts.

### 1.6. Randomization

The two treatment conditions (active TENS or sham TENS) were allocated using a minimisation procedure with stratification adjustments for pain intensity (0-10 pain scale), fibromyalgia severity (i.e. FIQ), and age. To optimize group balance, minimisation weights of 3 were assigned to pain intensity and fibromyalgia severity, and a weight of 1 was assigned to age. Allocation via minimisation was managed using Minimpy software (version 2.0.) on a password-protected computer. One of the TENS operators was involved in both participant enrolment and assignment via minimisation. The TENS operators were the only ones to have access to the allocation, which was performed after completion of pre-visit questionnaires. This procedure is considered a valid equivalent to conventional randomisation and could even be preferable for studies with small sample size according to CONSORT recommendations (29).

### 1.7. Blinding

To ensure blinding, participants were not directly informed about the existence of a sham stimulation condition until after data collection was completed. Participants were told that there were two different types of active TENS stimulation, which might or might not produce sensations, and that the presence or absence of sensations do not affect treatment effectiveness (26). Participant consent was reconfirmed during debriefing, when the sham condition was disclosed. The TENS unit used for sham was identical to the real intervention device but had modified cables preventing current delivery.

The TENS operator, responsible for preparing the TENS unit prior to the visit, delivering the TENS intervention and collecting participant expectation data, was not involved in outcome assessment or data analysis. The participant remained in the same room throughout the visit. At the very beginning of the visit, both the TENS operator and the evaluator were present together to reconfirm eligibility criteria. After this initial step, the two research staff members alternated participant interaction in separate rooms to maintain blinding, isolating the assessor during TENS procedures and the operator during assessments, each using headphones when absent. The TENS operator covered the electrodes with taped gauze to conceal any potential muscle contractions caused by the stimulations and instructed participants to clip the TENS device onto the hip opposite to where the outcome assessor was positioned. Participants were also explicitly instructed not to disclose any sensations or details related to the TENS intervention to the outcome assessor. The outcome assessor had no access to allocation lists, participant expectation responses, or pre-questionnaire data throughout the study.

Because expectations can strongly influence analgesic effect of TENS (26), participants expectations regarding treatment were assessed both before and after the first 10 minutes of TENS application using a scale from -100 (treatment will worsen pain) to +100 (treatment will completely relieve pain). This approach also allowed for indirect evaluation of blinding success, as revealing the presence of a sham group could have compromised blinding.

Blinding success was also evaluated by asking the evaluator to identify each participant’s arm allocation after trial completion. The evaluator, who was also in charge of completing data analysis, remained blinded to arm allocation until the statistical analysis was completed. The TENS operator, responsible for delivering the TENS intervention and collecting participant expectation data, was not involved in outcome assessment and data analysis.

### 1.8. Statistical Methods

Descriptive data are presented as mean *±* standard deviation (SD) and median (min-max) for continuous variables, and number (percent) for categorical variables. Between-group comparisons were performed using Mann-Whitney U-tests for continuous variables and Fisher’s exact or Chi-square tests for categorical variables. Within-group changes over time (compared to T1) were assessed with Friedman tests, and Kruskal-Wallis tests were used for between-subjects’ comparisons. Blinding success was evaluated by comparing participant expectations before and after the first 10 minutes of treatment using the Wilcoxon signed-rank test and by testing the outcome assessor’s guessing accuracy against chance (50%) with a binomial test. A responder analyses was also conducted for significant change scores to determine individual treatment responses. Finally, additional correlational and between-group exploratory analyses were conducted to look for potential effects of the TENS operator, leg press knee angle, caffeine consumption and kinesiophobia. Effect sizes were reported for pain outcomes in accordance to Tomczak and Tomczak’s recommendations (30) : Kendall’s W (ES_W_) was calculated for Friedman test, the coefficient r (ES_r_) was calculated for the Mann-Whitney U-test and for Friedman’s post-hoc (Wilcoxon signed-rank test), and Phi (ESφ) was calculated for Chi-square tests. To enhance reproducibility and reduce false positives, statistical significance was set at p<0.005, while p values between 0.05 and 0.005 were considered “suggestive”, as proposed by Benjamin et al. (31). All analyses were conducted using SPSS (IBM Corp, Armonk, NY, USA).

## Results

Participants were recruited between February 18 and June 27, 2025. The trial was completed on July 10, 2025. All participants who completed the experimental visit were included in the analysis. Three participants (2 from the active group and 1 from the sham group) did not attend the experimental visit (see figure 2) and were excluded from the analysis. Gender identity data were collected separately from biological sex. All participants were assigned female at birth. Of these, 20 identified as women, and one participant in the sham condition identified as a man. No participants identified as non-binary or other gender identities.

**Figure 2.**
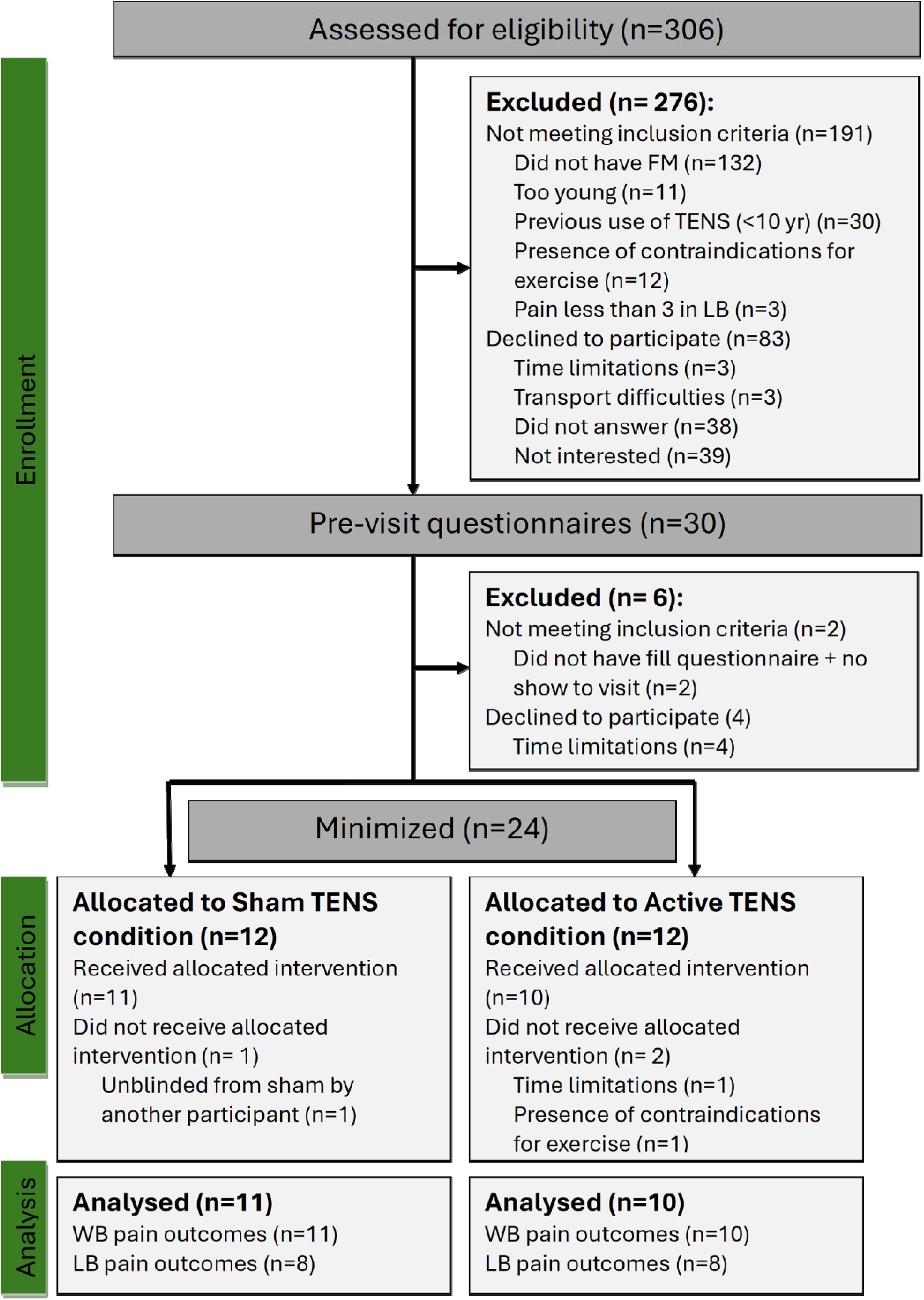
CONSORT flow diagram (29). WB Whole-body; LB Lower Back and Lower Body

**Table 1** summarizes participants characteristics by intervention condition. No statistically suggestive or significant differences were found between conditions for any of the variables (all p-values>.05). While the active TENS group showed numerically lower FIQ scores and higher activity levels, these differences were not statistically meaningful, and both groups were classified as having moderate fibromyalgia severity (32). The exercise intervention, as well as the TENS intervention and sham comparator were delivered as described in the Methods section by trained personnel. No adverse events were observed during exercise and TENS application.

**Table 1.**
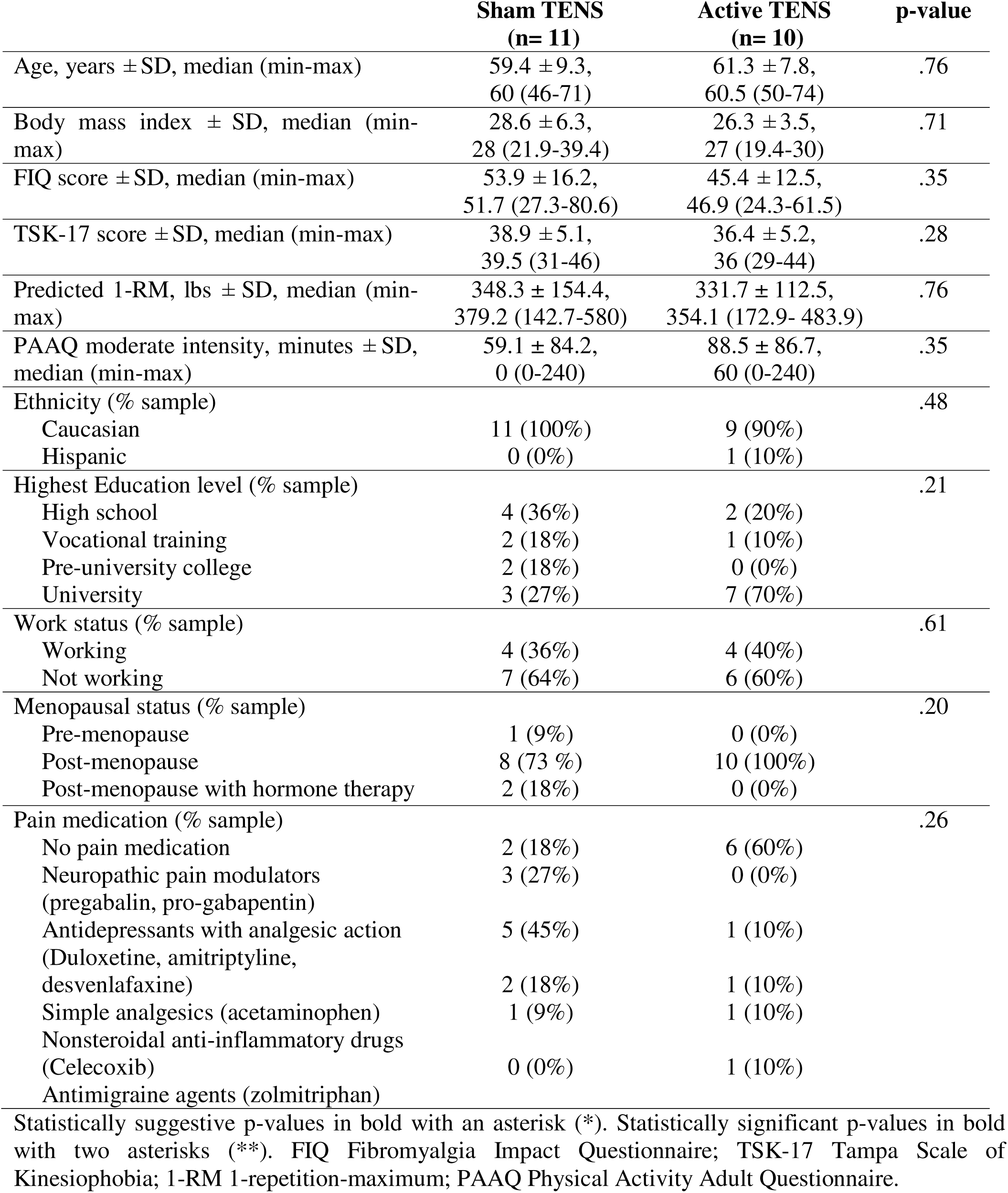
Participants characteristics.

### 1.9. Pain outcomes

#### 1.9.1. Whole body pain intensity

No statistically suggestive or significant differences in WB pain intensity over time were observed for the sham condition (p=.07, ES_W_=0.20). In contrast, the active condition revealed statistically meaninful differences between T1 and T2 (p =.048, ES_r_=0.63), as well as between T1 and T3 (p=0.004, ES_r_=0.92). The other pairwise comparisons are presented in table S1.

As for between-condition comparisons, statistically significant differences in WB pain intensity were observed in the active condition compared to the sham condition at T2 in favor of the active condition (p=.001, ES_r_=0.70). Similarly, at T3, WB pain intensity was significantly lower in the active condition compared to the sham condition (p <.001, ES_r_=0.78). No statistically suggestive or significant differences between conditions were found at T1 (baseline), T4 or T5 (table 2).

**Table 2.** Pain outcomes over time by intervention condition.

| Measures | Time point | Whole-Body |  |  | Lower Back and Lower Body |  |  |
| --- | --- | --- | --- | --- | --- | --- | --- |
| | | Sham TENS<br>Mean ± SD<br>Median<br>(min-max) | Active TENS<br>Mean ± SD<br>Median<br>(min-max) | Between-Group<br>Effect Size (r) | Sham TENS<br>Mean ± SD<br>Median<br>(min-max) | Active TENS<br>Mean ± SD<br>Median<br>(min-max) | Between-Group<br>Effect Size (r) |
| Pain Intensity | T1 | 6.3 ± 1.4<br>6.0 (4-8) | 5.0 ± 1.9<br>5.0 (2-8) | 0.34 | 6.5 ± 0.5<br>6.5 (6-7) | **3.38 ± 2.3\***<br>**3.0 (0-7)** | 0.65 |
| | T2 | 7.36 ± 0.9<br>7.0 (6-9) | **4.1 ± 2.3\*\***<br>**4.0 (1-7)** | 0.70 | 7.4 ± 1.5<br>8.0 (4-9) | **4.4 ± 2.5\***<br>**5.0 (1-8)** | 0.61 |
| | T3 | 6.8 ± 1.5<br>7.0 (4-9) | **3.3 ± 1.4\*\***<br>**3.5 (1-6)** | 0.78 | 5.9 ± 2.2<br>6.0 (2-9) | **3.4 ± 1.4\***<br>**3.5 (1-5)** | 0.57 |
| | T4 | 6.3 ± 3.0<br>7.0 (1-10) | 4.8 ± 2.3<br>5.0 (2-8) | 0.37 | 6.3 ± 2.0<br>7.0 (2-8) | 4.5 ± 2.3<br>5.0 (1-8) | 0.41 |
| | T5 | 5.8 ± 2.2<br>6.0 (1-9) | 4.9 ± 2.4<br>5.0 (1-8) | 0.18 | 5.1 ± 1.9<br>5.5 (3-7) | 3.4 ± 2.6<br>3.0 (1-8) | 0.36 |
| Pain Unpleasantness | T1 | 6.3 ± 1.8<br>6.0 (4-10) | 5.2 ± 2.4<br>6.0 (0-8) | 0.15 | 6.3 ± 0.7<br>6.0 (5-7) | **4.0 ± 2.8\***<br>**4.0 (0-8)** | 0.41 |
| | T2 | 7.1 ± 1.4<br>8.0 (5-9) | **3.8 ± 2.4\*\***<br>**3.5 (0-7)** | 0.66 | 7.4 ± 1.3<br>8.0 (5-9) | **4.2 ± 2.1\*\***<br>**4.5 (0-7)** | 0.72 |
| | T3 | 6.1 ± 2.6<br>7.0 (0-9) | **3.3 ± 1.8\*\***<br>**3.0 (0-7)** | 0.60 | 5.6 ± 2.4<br>5.5 (2-9) | **3.3 ± 1.4\***<br>**4.0 (0-4)** | 0.51 |
| | T4 | 6.2 ± 2.5<br>6.0 (2-10) | 4.3 ± 2.5<br>4.5 (0-8) | 0.37 | 5.8 ± 2.4<br>6.5 (3-8) | 4.2 ± 2.4<br>3.5 (1-9) | 0.29 |
| | T5 | 5.9 ± 2.2<br>6.0 (1-9) | 4.7 ± 2.9<br>5.0 (0-8) | 0.21 | 5.5 ± 2.3<br>6.0 (2-8) | 3.9 ± 2.8<br>4.0 (0-8) | 0.29 |

Pain was measured on 0-10 numerical scales; pain intensity: 0=no pain, 10=unbearable pain; pain unpleasantness: 0=not unpleasant, 10=extremely unpleasant. T1: baseline; T2: immediately post-exercise; T3: 15 minutes post-exercise; T4: end of the day following visit; T5 24 hours post-exercise. Statistical suggestive p-value ( $p < .05$ ) are in bold with an asterisk (\*) and significant p-value ( $p < .005$ ) are in bold with two asterisks (\*\*) to indicate inter-condition analyses. The coefficient r was calculated to report effect sizes.

Between-group comparisons for delta scores showed a statistically suggestive difference for ΔT2–T1 (p=.01, ES_r_=0.57) and a statistically significant difference for ΔT3–T1 (p=.001, ES_r_=0.70), respectively. No statistically suggestive differences in WB pain intensity were found between conditions for ΔT4–T1 (p=.71, ES_r_=0.09) or ΔT5–T1 (p=.76, ES_r_=0.08; see figure 3A).

**Figure 3.**
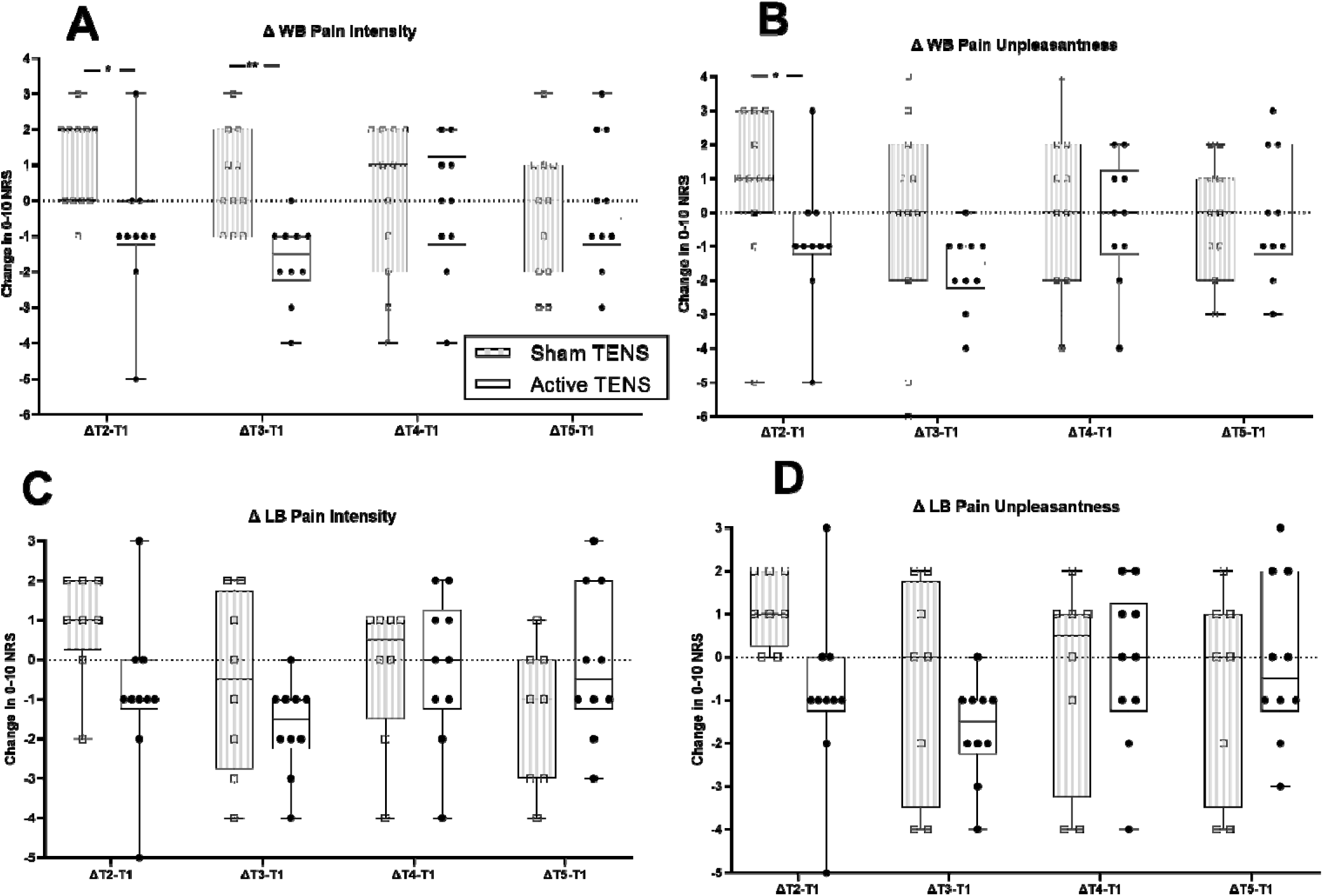
Changes in pain intensity and unpleasantness scores over time by TENS condition. Panels A and B show changes in whole-body (WB) pain intensity (A) and pain unpleasantness (B), while panels C and D show changes in low-back and lower body (LB) pain intensity (C) and pain unpleasantness (D), measured on a 0–10 numerical rating scale (NRS). Data represent change scores from baseline (T1) to subsequent time points (T2 to T5). Boxplots display medians, interquartile ranges, and individual data points. Positive values indicate increased pain or unpleasantness relative to baseline, while negative values indicate decreases. Statistical significance of between-cond tion differences is indicated by asterisks: *p<.05, **p<.005.

#### 1.9.2. Whole body pain unpleasantness

No statistically suggestive or significant differences in WB pain unpleasantness over time were observed for the sham condition (p=.06, ES_w_=0.21). In the active condition, statistically suggestive differences were observed between T1 and T2 (p=.01, ES_r_=0.78), and a statistically significant decrease was found between T1 and T3 (p<.005, ES_r_=0.89). The other pairwise comparison are presented in table S1.

For between-condition comparisons, WB pain unpleasantness was significantly lower in the active condition compared to the sham condition at T2 (p=.002, ES_r_=0.66) and T3 (p <.005, ES_r_=0.60). No statistically suggestive or significant differences were found between conditions at T1, T4 and T5 (table e 2).

Between-group comparisons were statistically suggestive for ΔT2–T1, with the sham group showing an increase in pain and the active group a decrease in pain (p=.006, ES_r_=0.59). No suggestive differences in WB pain unpleasantness were found between conditions for ΔT3–T1 (p=.06, ES_r_=0.42), ΔT4–T1 (p=.39, ES_r_=0.19) or ΔT5–T1 (p=.86, ES_r_=0.05) (figure 3B).

#### 1.9.3. Low back and lower limb pain intensity

A statistically suggestive *increase* in LB pain intensity were observed in the sham condition between T1 and T2 (p=.048, ES_r_=0.70). All comparisons are presented in table S1. Between-condition comparisons revealed a statistically suggestive difference between the active and sham condition at T1 (p=.01, ES_r=_0.65), T2 (p=.01, ES_r=_0.61) and T3 (p=.02, ES_r=_0.57), favoring the active condition. Between-condition comparisons at T4 and T5 revealed no statistically and suggestive or significant differences (table 2). No between-group comparisons of the Δ values showed statistically suggestive or significant differences (all p>.20, all ES_r_<0.33; figure 3C).

#### 1.9.4. Lower back and lower limb pain unpleasantness

Between-condition comparisons showed a significantly lower LB pain unpleasantness in the active condition at T2 (p=.003, ES_r=_0.72) and a suggestively lower LB pain unpleasantness at T3 (p =.048, ES_r_=0.51). No suggestive differences were observed at T4 or T5 (table 2). No statistically suggestive or significant between-group differences were observed for delta scores (all p>0.28, all ES_r_<0.28; figure 3D).

#### 1.9.5. Responders to TENS intervention

TENS responders were defined as participants who achieved the minimal clinically important difference, operationalized as either a reduction of at least 1 point or 15% (minimal change) on the 0– 10 NRS, or a reduction of at least 2 points or 33% on the NRS, the latter being associated with a rating of “much better” improvement (33,34).

Responder rates are presented in table 3. Between T2 and T1, a significant higher proportion of active TENS participants achieved ≥15% and ≥33% reduction in WB pain intensity compared to sham. This difference was even more pronounced between T3 and T1, with 90% of active TENS participants achieving a ≥15% reduction and 60% achieving ≥33% reduction, versus none in the sham condition. For WB unpleasantness, a statistically suggestive differences favoring the active condition was observed between T2 and T1 for the ≥15% threshold. No statistically suggestive differences were found at other thresholds or time points.

**Table 3.**
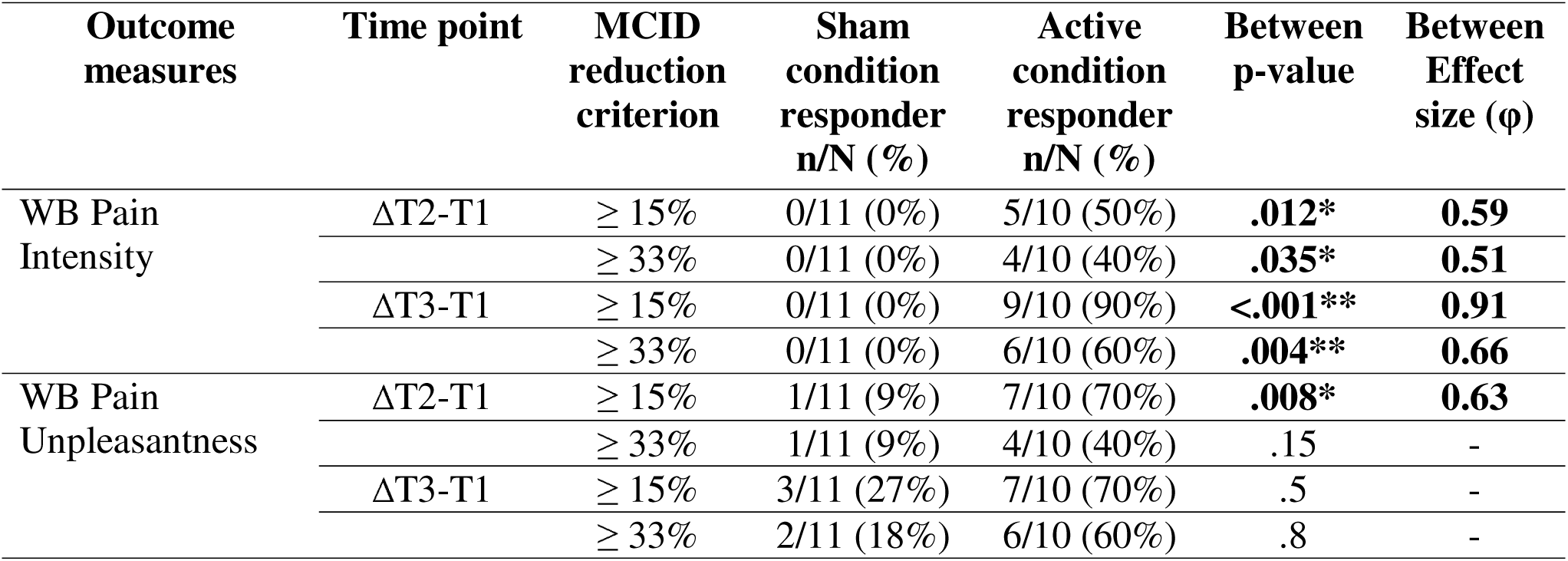
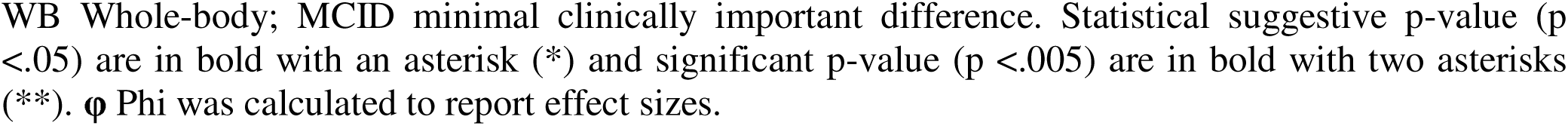
Responder rates based on minimal clinically important difference criteria for pain outcomes.

| Outcome measures | Time point | MCID reduction criterion | Sham condition responder n/N (%) | Active condition responder n/N (%) | Between p-value | Between Effect size ($\phi$) |
| --- | --- | --- | --- | --- | --- | --- |
| WB Pain Intensity | ΔT2-T1 | $\geq 15\%$ | 0/11 (0%) | 5/10 (50%) | **.012\*** | **0.59** |
| | | $\geq 33\%$ | 0/11 (0%) | 4/10 (40%) | **.035\*** | **0.51** |
| | ΔT3-T1 | $\geq 15\%$ | 0/11 (0%) | 9/10 (90%) | **<.001\*\*** | **0.91** |
| | | $\geq 33\%$ | 0/11 (0%) | 6/10 (60%) | **.004\*\*** | **0.66** |
| WB Pain Unpleasantness | ΔT2-T1 | $\geq 15\%$ | 1/11 (9%) | 7/10 (70%) | **.008\*** | **0.63** |
| | | $\geq 33\%$ | 1/11 (9%) | 4/10 (40%) | .15 | - |
| | ΔT3-T1 | $\geq 15\%$ | 3/11 (27%) | 7/10 (70%) | .5 | - |
| | | $\geq 33\%$ | 2/11 (18%) | 6/10 (60%) | .8 | - |

WB Whole-body; MCID minimal clinically important difference. Statistical suggestive p-value ( $p < .05$ ) are in bold with an asterisk (\*) and significant p-value ( $p < .005$ ) are in bold with two asterisks (\*\*). $\phi$ Phi was calculated to report effect sizes.

### 1.10. Participants Expectations

There were no statistically significant or suggestive differences in treatment expectations between the active and sham (p=.92) groups, both before and after 10 minutes of TENS application (p=.76), indicating comparable expectations regarding TENS effect and supporting the adequacy of blinding.

### 1.11. Outcome Assessor Blinding

The assessor correctly identified the condition (active vs. sham TENS) in 14 out of 21 cases. Such performance (67%) did not differ from chance (50%; p=.19), suggesting that assessor blinding was maintained.

### 1.12. TENS Operator

Given that the intervention was administered by two individuals of different sexes (one female [n=11], one male [n=10]), exploratory analyses were conducted to assess whether the TENS operator influenced the outcomes. Both the sham and active TENS conditions showed greater reduction in WB pain unpleasantness between T2 and T1 and between T3 and T1 for the female TENS operator compared to the male TENS operator, reaching statistical significance for the sham TENS condition (p<.009) and showing a statistically suggestive trend for the active TENS condition (p<.04, see figure S2). A similar pattern of results was observed for LB pain unpleasantness for the active TENS condition between T2 and T1 (p=.04), as well as between T3 and T1 (p=.04), suggesting greater TENS effect on pain unpleasantness with the female operator.

### 1.13. Leg press knee angle

Given the potential impact of exercise demands and biomechanics on pain outcomes, and the fact that 6 (out of 21) participants were unable to complete the exercise at a 90-degree knee angle, we explored whether knee angle during exercise influenced pain outcomes. Results revealed that, for the sham condition, participants exercising at 90° showed a statistically suggestive greater decrease in LB pain intensity between T1 and T5 (–2.75 ± 1.58) compared to those at 120° (0.0 ± 0.82, p=.03). No other suggestive or statistically significant effects were observed in either the sham or active conditions.

### 1.14. Caffeine

To minimize potential confounding effects, participants were instructed to abstain from caffeine consumption for at least 6 hours prior to the exercise session conducted at the research center (35). Post-session intake, however, was more difficult to control once participants returned home. Accordingly, Spearman exploratory analyses were conducted to investigate potential associations between home pain outcomes (T4 and T5) and caffeine consumption.

Analyses were conducted for both the total amount of caffeine (mg) and the time of consumption. The later was operationalized as a categorical variable, taking into account the half-life of the molecule (36). The variable was coded as follow: 0=no caffeine intake, 1=caffeine consumed more than 6 hours before home TENS session, 2=caffeine consumed less than 6 hours before home TENS session.

For the active TENS condition, a suggestive and positive correlation was observed between the increase in WB pain intensity from T1 to T4 and both the total amount of caffeine consumed in mg on the evening of the session (r=0.71, p=.02) and more recent caffeine consumption (r=0.76, p=.01; see figure S3A). Recent caffeine consumption also suggestively correlated with increased LB pain intensity between T4 and T1 (r=0.72, p=.046; see figure S3B). No other suggestive correlations between caffeine intake and TENS effect were observed, including for the sham condition (all p-values >.06).

### 1.15. Kinesiophobia

Given that pre-exercise levels of kinesiophobia can influence exercise-induced pain intensity (37,38), we conducted exploratory analyses to examine the relations between baseline kinesiophobia and pain outcomes. In the sham condition, there were no statistically suggestive or significant correlations between the TSK-17 score and any change in pain. In contrast, for the active condition, higher TSK-17 scores were associated with greater reductions in LB pain intensity. Specifically, TSK-17 scores were suggestively and negatively correlated with changes in LB pain intensity from T1 to T4 (r=–0.75, p=.03) and from T1 to T5 (r=–0.83, p=.01), as well as with changes in LB pain unpleasantness from T1 to T4 (r=–0.78, p=.02) and from T1 to T5 (r=–0.79, p=.02). No additional statistically suggestive or significant correlations were observed in the active condition.

## Discussion

The aim of this randomized controlled trial was to investigate the effect of a single session of conventional TENS applied during and in the day following a leg press exercise on short-term MEP intensity and unpleasantness in women with fibromyalgia. Our results reveal that a single session of conventional TENS, applied before and during a lower-body resistance exercise, can reduce pain intensity and unpleasantness immediately and 15 minutes after exercise in women with fibromyalgia, although these effects diminish by the evening and 24-h post-exercise. In the early window, the observed benefits were greater than sham stimulation, suggesting effects beyond placebo. Improvements were most evident at 15 minutes after exercise, consistent with the early phase when MEP could be the strongest (10,39,40). To our knowledge, this is the first study to evaluate TENS effects on MEP following resistance exercise in women with fibromyalgia, addressing a critical gap given resistance training’s delayed symptomatic improvement (8).

Assessment of clinically meaningful change revealed that 40% of individuals in the active TENS cohort achieved ≥33% pain reduction at T2, with the proportion increasing to 60% at T3 (34). These proportions are consistent with findings by Dailey et al. (16), who reported that 44% of women with fibromyalgia experienced more than 30% pain reduction during functional tasks with active TENS. In their study, Dailey et al. (16) investigated the acute effect of a single high-frequency TENS session during functional tests (6-minute walk test [6MWT] and 5-sit-to-stand test [5-STS]) in women with fibromyalgia, reporting mean pain reductions of -1.8 and -1.6 points, respectively. In our study, which examined the effects of a single high-frequency TENS session administered before and during a resistance exercise session, we observed a smaller immediate post-exercise pain reduction of -0.9 point that increased to -1.7 points 15-minutes later. These findings closely align with those of Dailey et al. (16), suggesting comparable hypoalgesic effect across different physical task contexts. The smaller reductions observed immediately after exercise in our study could reflect the greater intensity of resistance exercise compared to functional tests, as MEP is known to depend on movement type, intensity, and duration (10).

Importantly, group differences were no longer evident at the later assessments, at the end of the day and 24 hours post-exercise. A closer examination of the data reveals a trend toward stable or slightly reduced pain scores in the sham group, whereas the active TENS group showed a tendency for increased pain relative to immediately post-exercise (T2) (see again figure 4). This pattern may reflect a waning of the immediate effect of TENS, the impact of potential confounders, such as caffeine use after leaving the lab, but also the natural decrease in MEP in the sham condition.

Caffeine intake, uncontrolled beyond the 6 hour pre visit restriction, showed associations with greater subsequent pain intensity in active condition, in line with evidence that caffeine can interfere with high frequency TENS analgesia (35). Psychological and interpersonal factors also emerged. In the active condition of our study, higher kinesiophobia was unexpectedly associated with reduced lower-body pain at later time points (37), potentially reflecting behavioral adaptations such as activity pacing or reduced exertion, leading to lower effective exercise intensity. Although all participants exercised at 60% of predicted 1-RM, greater kinesiophobia could have biased the strength estimate and lowered the actual exercise intensity. Given that movement-evoked pain varies with task type, intensity, and context (10), exercising at a lower effective intensity may explain the negative association observed. From a clinical standpoint, these findings may indicate that individuals with the highest levels of kinesiophobia are also those most likely to benefit from TENS, with the intervention being particularly effective in reducing post-exercise pain sensitivity in this population. Additional research is required to validate these preliminary results.

Exploratory analyses also suggested that knee angle during the leg press may influence later pain responses. In the sham condition, participants exercising at 90° knee flexion showed a greater reduction in LB pain intensity at 24-h compared to those at 120°. This is somewhat surprising, as deeper knee flexion is typically associated with increased gluteus maximus and quadriceps activation (39), which would be expected to increase MEP since it’s known to increase exercise induced hypoalgesia in healthy population (42). However, this effect was not observed in the active TENS group, raising the possibility of a type I error for the sham group. Alternatively, the unequal subgroup sizes and potential inter-individual variability may have contributed to a type II error in the active group, again limiting the overall interpretation of these findings.

Additionally, pain outcomes measured at the research center (T2 and T3) differed based on operator sex: participants attended by a female operator reported greater reductions in pain unpleasantness regardless of condition, consistent with evidence that experimenter characteristics and nonverbal cues can influence pain perception and placebo responses (40). These results highlight the role that both behavioral and contextual factors can play in shaping MEP responses and TENS effects.

Directly measuring blinding by asking participants to guess their treatment allocation or whether their treatment was “real” (26) could have led them to suspect the presence of a placebo condition, potentially biasing their responses and potentially undermining the blinding procedure. Additionally, expectations can significantly influence the analgesic effect of TENS (26). Thus, changes in participants’ treatment expectations were analysed as a proxy for blinding effectiveness and to explore the potential influence of expectations regarding TENS on outcome measures. There were differences between the active and sham condition for participants’ treatment expectations, either before treatment or after the first 10 minutes of TENS intervention, suggesting effective blinding and indicating that expectations about TENS did not account for group differences reported in this study. Assessor blinding was also maintained, although the proportion of correct guesses by the assessor was higher than chance. While this difference was not statistically significant, caution is warranted given the relatively small sample size and the chances of committing a type II error.

This study contributes novel evidence on the acute potential of conventional TENS on MEP following resistance training in women with fibromyalgia. The double-blind design strengthens the internal validity of the findings, and the inclusion of multiple post-exercise time points allows a nuanced and comprehensive understanding of both immediate and “longer-term” effects. Conversely, certain limitations should also be acknowledged. For example, the sample size, while adequate for detecting acute effects (main outcome), may have limited statistical power to detect smaller or subgroup effects, such as differences at later time points (T4 and T5). Caffeine consumption was controlled prior to the experimental visit, but their intake was self-reported and not controlled experimentally once participants returned home. The study population was limited to women with fibromyalgia, which improves internal consistency but may reduce generalizability to men or other chronic pain populations.

## Conclusion

Together, these findings provide the first evidence that a single session of conventional TENS applied during resistance training may help reduce MEP intensity and unpleasantness in women with fibromyalgia, potentially easing an important obstacle to exercise. Although these effects were not sustained over time — likely due to external influences and limited statistical power — these findings suggest that TENS can help mitigate acute post-exercise pain. Future research with larger cohorts is needed to confirm these results, evaluate the effects of repeated applications, and explore personalized TENS dosing to optimize outcomes in clinical practice. Furthermore, by alleviating acute post-exercise pain, this intervention warrants rigorous investigation as a promising adjunct to facilitate long-term adherence to resistance exercise programs.

## Acknowledgement

Funding: MR received a Master’s scholarship from the Canadian Institutes of Health Research (CIHR/IRSC) [2023] and the Fonds de recherche du Québec – Santé (FRQS) [2023-2025]; GL received salary support from the Fonds de recherche du Québec – Santé (FRQS) [2022-2025]. This research was made possible thanks to a testamentary donation designated for fibromyalgia research [2021].

## CRediT Roles

**Mathil Ruel:** Conceptualization, Methodology, Software, Investigation, Data Curation, Formal analysis, Visualization, Project administration, Writing - Original Draft, Writing - Review & Editing; **Shayana Lussier Crevier**: Software, Resources, Investigation, Data Curation, Writing - Review & Editing; **Nicolas Dehors:** Investigation; **Serge Marchand:** Funding acquisition, Writing - Review & Editing; **Isabelle Dionne:** Resources, Supervision, Writing - Review & Editing; **Guillaume Léonard:** Methodology, Resources, Supervision, Writing - Review & Editing

## Data availability

All data are available upon reasonable request to the corresponding author.

