## Supplementary for "TENS for Movement-Evoked Pain in Fibromyalgia: A Randomised Double-Blind Controlled Trial"

Supplementary materials

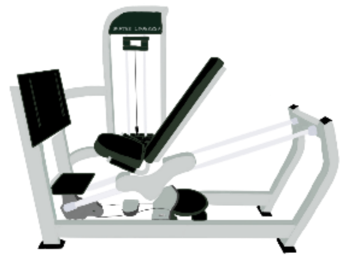

**Supplementary Figure 1. Seated Leg Press Machine** © Mathil Ruel, 2025

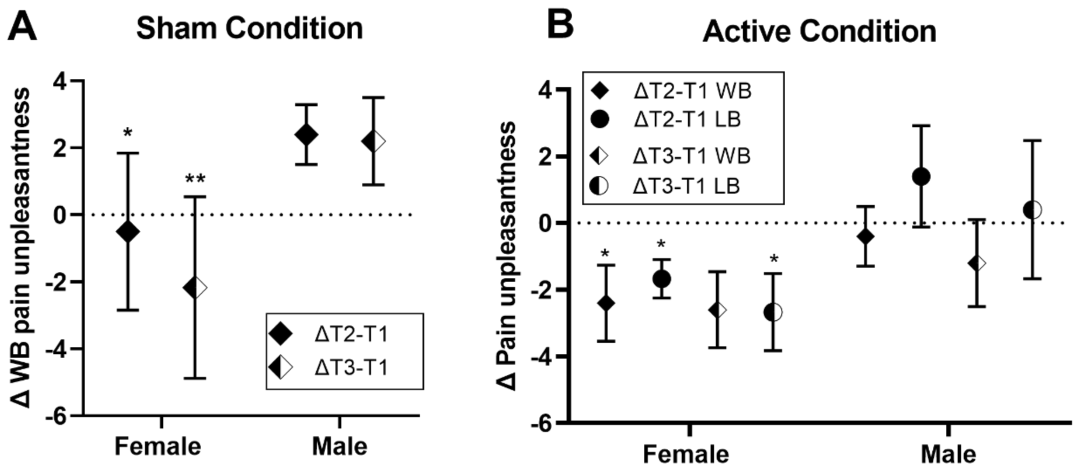


**Supplementary Figure 2.** Effect of TENS administrator on changes in pain unpleasantness in active and sham conditions. (A) Change in pain unpleasantness (delta scores) from baseline (T1) to T2 and T3 for whole-body (WB) in the sham TENS condition, separated by administrator sex (female vs. male). (B) Change in pain unpleasantness (delta scores) from baseline to T2 and T3 for WB and lower body and lower back (LB) regions in the active in the sham condition, separated by administrator sex. Significant between-operator sex differences are indicated by asterisks (*p < .05, **p < .005).

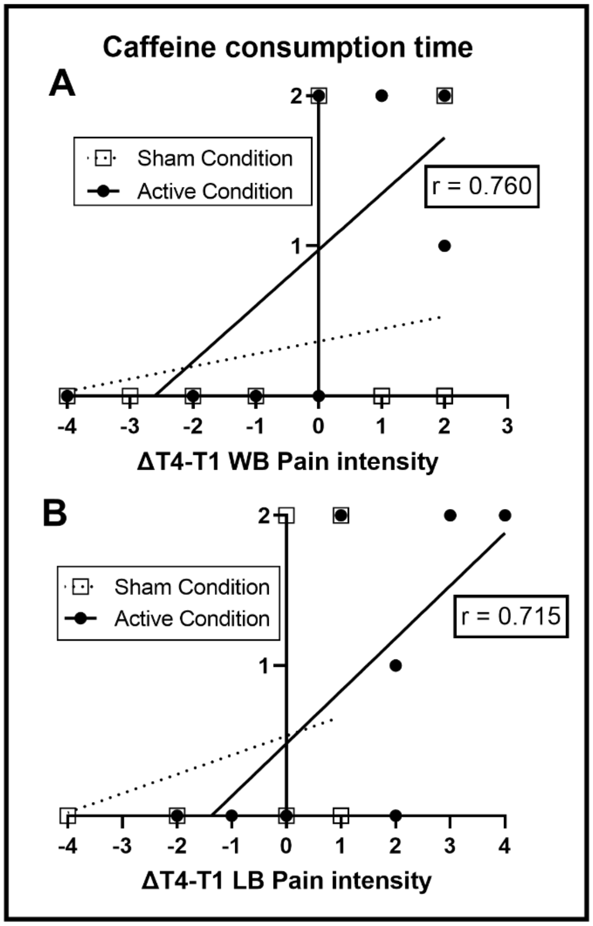


**Supplementary Figure 3.** Association Between Caffeine Consumption Timing and Pain Intensity Change. Timing of caffeine consumption coded categorically as 0 = no caffeine intake, 1 = caffeine consumed more than 6 hours before TENS session, 2 = caffeine consumed less than 6 hours before TENS session. Panel A displays whole-body (WB) pain intensity change between T1 and T4, and Panel B displays lower-body (LB) pain intensity change between T1 and T4. A suggestive strong positive correlation (active: $r = 0.760$ for WB pain intensity, $r = 0.715$ for LB pain intensity, $p < .05$) was observed, where more recent caffeine consumption is associated with increased pain intensity in the active group.

**Supplementary Table 1.** Within-group changes in pain intensity and unpleasantness over time by intervention condition.

| Measures | Time comparison | Whole-body | | | | | | Lower body | | | | | |
|---|---|---|---|---|---|---|---|---|---|---|---|---|---|
| | | Within Sham TENS | | | Within Active TENS | | | Within Sham TENS | | | Within Active TENS | | |
| | | Δ-value | p | Effect Size | Δ-value | p | Effect Size | Δ-value | p | Effect Size | Δ-value | p | Effect Size |
| **Pain Intensity** | **T1-T2** | -1.1 | .07 | 0.20 | 0.9 | **.048*** | 0.63 | -0.9 | **.048*** | 0.70 | -1.02 | .48 | 0.11 |
| | **T1-T3** | -0.5 | - | | 1.7 | **.004**** | 0.92 | 0.6 | .81 | 0.08 | -0.02 | - | |
| | **T1-T4** | 0.0 | - | | 0.2 | .83 | 0.07 | 0.2 | .94 | 0.03 | -1.12 | - | |
| | **T1-T5** | 0.5 | - | | 0.1 | .36 | 0.29 | 1.4 | .30 | 0.36 | -0.02 | - | |
| | **T2-T3** | 0.6 | - | | 0.8 | .36 | 0.29 | 1.5 | **.03*** | 0.78 | 1 | - | |
| | **T2-T4** | 1.1 | - | | -0.7 | .08 | 0.56 | 1.1 | .058 | 0.67 | -0.1 | - | |
| | **T2-T5** | 1.6 | - | | -0.8 | .29 | 0.34 | 2.3 | **.003**** | 1.00 | 1 | - | |
| | **T3-T4** | 0.5 | - | | -1.5 | **.007*** | 0.85 | -0.4 | .75 | 0.11 | -1.1 | - | |
| | **T3-T5** | 1.0 | - | | -1.6 | **.048*** | 0.63 | 0.8 | .43 | 0.28 | 0 | - | |
| | **T4-T5** | 0.5 | - | | -0.1 | .48 | 0.22 | 1.2 | .27 | 0.39 | 1.1 | - | |
| **Pain Unpleasantness** | **T1-T2** | -0.8 | .06 | 0.21 | 1.4 | .01 | 0.78 | -1.1 | .10 | 0.24 | -0.2 | .34 | 0.14 |
| | **T1-T3** | 0.2 | - | | 1.9 | **.0047**** | 0.89 | 0.7 | - | | 0.7 | - | |
| | **T1-T4** | 0.1 | - | | 0.9 | .44 | 0.25 | 0.5 | - | | -0.2 | - | |
| | **T1-T5** | 0.4 | - | | 0.5 | .32 | 0.31 | 0.8 | - | | 0.1 | - | |
| | **T2-T3** | 1.0 | - | | 0.5 | .72 | 0.11 | 1.8 | - | | 0.9 | - | |
| | **T2-T4** | 0.9 | - | | -0.5 | .09 | 0.54 | 1.6 | - | | 0 | - | |
| | **T2-T5** | 1.2 | - | | -0.9 | .14 | 0.47 | 1.9 | - | | 0.3 | - | |
| | **T3-T4** | -0.1 | - | | -1.0 | **.04*** | 0.65 | -0.2 | - | | -0.9 | - | |
| | **T3-T5** | 0.2 | - | | -1.4 | .07 | 0.58 | 0.1 | - | | -0.6 | - | |
| | **T4-T5** | 0.3 | - | | -0.4 | .83 | 0.07 | 0.3 | - | | 0.3 | - | |

Mean differences (Δ) in pain intensity and unpleasantness scores between specified time points are shown for whole-body and lower-body regions in active and sham TENS conditions. Negative values indicate increases in pain or unpleasantness over time. Statistical suggestive p-value ($p < .05$) are in bold with an asterisk (*) and significant p-value ($p < .005$) are in bold with two asterisks (**). Kendall's W was used to report effect sizes of Friedman tests, and the coefficient r was used for Wilcoxon signed-rank tests (post-hoc).
